# Establishing effective partnerships among case managers and clinicians to improve linkage to and engagement in HIV care: A scoping review

**DOI:** 10.1101/2025.05.10.25327371

**Authors:** Piring’ar Mercy Niyang, Ibrahim Bola Gobir, Itunu Dave-Agboola, Joy Musa Shallangwa, Muhammad Zaharaddeen, Bashir Zubayr, Chika Obiora-Okafor, Terso Usha, Mabel Ikpeme, Onyeka Igboelina, Pamela Gado, Kolawole Olatunbosun, Dolapo Ogundehin

**Affiliations:** Georgetown Global Health Nigeria; United States Agency for International Development

**Keywords:** Care continuum, antiretroviral therapy, clinician-case manager partnerships, retention in care, linkage to care, patient-centered care, interdisciplinary collaboration, psychosocial support, stigma, Sub-Saharan Africa, barriers to care, holistic care models, community-based programs, people living with HIV

## Abstract

HIV continues to pose significant health challenges globally, particularly in resource-limited settings such as Sub-Saharan Africa, which bears the brunt of the epidemic. The HIV care continuum, encompassing testing, linkage to care, retention, and antiretroviral treatment (ART) adherence, is critical in reducing morbidity and mortality while improving the quality of life for people living with HIV (PLHIV). Despite its potential, key HIV care continuum events such as linkage to treatment, retention in care and adherence remain substantial challenges due to barriers such as stigma, socio-economic inequalities, and systemic inefficiencies. This scoping review explores effective partnerships between clinicians and case managers to address these challenges. Using a Joanna Briggs Institute framework and population, concept and context (PCC) guidelines, 31 studies were analyzed, highlighting successful collaborative models, barriers, and facilitators in implementing effective HIV care. Results gotten from the review highlight the importance of role clarity, interdisciplinary approaches and collaborations, and patient-centered models integrating clinical and psychosocial support. Case managers, by addressing logistical challenges, complement clinicians’ medical expertise, resulting in improved ART adherence and retention. Effective interventions, such as holistic care models and community-based programs, were found to mitigate barriers like stigma and housing instability, and resource constraints. Recommendations call for enhanced training, clear role definitions, and policies fostering collaboration between case managers and clinicians to optimize care delivery. Strengthening these partnerships between clinicians and case managers is essential for the HIV care continuum and achieving better health outcomes for PLHIV.

## Background

HIV infection continues to pose a critical health challenge worldwide, particularly in low-income developing nations (Suleiman and Momo, 2016). According to the World Health Organization (WHO), over 85.6 million people have been infected with HIV, and more than 40.4 million HIV-related deaths have been recorded (World Health Organization, 2024). Notably, 71.0% of PLHIV reside in Sub-Saharan Africa, despite this region comprising just over 12% of the global population (Suleiman and Momo, 2016).

The HIV Continuum of Care (“care continuum”) encompasses HIV testing, linkage to care, retention and adherence to ART. This continuum has proven effective in reducing HIV transmission, morbidity, and mortality among those at risk or already infected and in improving treatment outcomes and quality of lives among PLHIV. Integrating HIV services with comprehensive primary healthcare has been recommended to improve health outcomes for individuals at higher risk of HIV exposure (Rahman et al., 2021).

Retention and adherence to ART remain significant challenges in managing HIV and care continuum. To address these issues, various strategies and interventions are being designed and implemented globally. HIV affects multiple facets of an individual’s life, including social, economic, physical, emotional, and spiritual well-being. PLHIV often face vulnerabilities such as poverty, stigma, discrimination, inequality, depression, guilt, prejudice, and anxiety, complicating efforts to maintain care retention and treatment adherence (Rachlis et al., 2016; Comins et al., 2024).

Multidisciplinary approaches are often necessary to address challenges associated with care continuum including adherence to ART and appointment-keeping, involving collaborations across nursing, pharmacy, social work, and case management. Clinicians in the context of this study refer to healthcare professionals such as physicians, nurse practitioners, and physician assistants, who provide direct medical care to individuals living with HIV (Khalili and Landovitz 2020). Their responsibilities include diagnosing HIV, initiating and managing ART, monitoring patients’ health status, and adjusting treatments as necessary to achieve viral suppression (Keene et al., 2023; Walker et al., 2023). They play a crucial role in helping patients understand the importance of adherence to the care continuum, identifying barriers to adherence, and addressing those within their capacity (Walker et al., 2023). Linking patients to resources, including case managers, is essential to overcoming additional barriers. Similarly, a Case Manger is typically a non-clinical professional or peer support worker who coordinates care services to ensure that individuals engage with and remain in the HIV care continuum (Besada et al., 2018). Case management support services are specialized services provided to HIV patients (Øgård-Repål et al., 2023). These services involve establishing strong relationships and communication between HIV service providers and their clients, ensuring efficient coordination of medical care and social support services (Besada et al., 2018; Keene et al., 2023). The ultimate goal of case management support services is to effectively engage and retain patients, ensuring good adherence to medication and overall treatment (Sarfo et al., 2017).

Effective partnership between the clinicians and case managers is essential for optimal HIV care continuum. Some factors may contribute to the prevention of this collaboration. E.g., unclear definitions of responsibilities between the two can lead to confusion and overlap, hindering teamwork, differences in communication styles and lack of standardized protocols can result in misinterpretations and fragmented care (Rahman et al., 2021). Limited staffing, high caseloads, difference in remuneration, lack of trust and inadequate training can strain the capacity of both case managers and clinicians, reducing opportunities for collaboration. High employee turnover among providers further exacerbates these challenges (Shamu et al., 2019; Fuller et al., 2024). Understanding the dynamics of these partnerships and identifying best practices will help improve the overall effectiveness of HIV care programs, thereby reducing transmission rates and improving health outcomes for PLHIV.

Despite Sub-Saharan Africa being the epicenter of new HIV infections, there is limited attention to care and support services for individuals with HIV/AIDS beyond ART programs (Mwai et al., 2013). This scoping review aims to explore best practices and successful models of collaboration between case managers and HIV clinicians, assess the impact of these partnerships on patient outcomes, particularly in terms of linkage to care and retention in HIV treatment, analyze the barriers and facilitators in forming effective partnerships between case managers and clinicians. Finally, the review will propose recommendations for policy and practice that can support the establishment and maintenance of these partnerships.

### Methodology

This scoping review was conducted using the Joanna Briggs Institute (2020) scoping review framework as described by Santos et al., (2018).

Literature search approach was based on “Population, Concept and Context” (PCC) guide as described by Peters et al., (2015).

**Table 1:** Literature search based on the PCC.

|  | Theme | Search Number | Search Terms |
| --- | --- | --- | --- |
| Population | People Living with HIV | S1 | “People Living with HIV” ,<br>HIV infected individuals, Key<br>populations, |
| Concept | Partnership between case<br>managers and HIV<br>clinicians | S2 | "case management," "clinician<br>partnership," “team-based<br>care”, “linkage to care”,<br>“retention in care” and<br>"treatment adherence." |
| Context | HIV care settings,<br>including clinics and<br>community health<br>organizations | S3 | "HIV care," and "patient<br>engagement," “community<br>health services”, “low- and<br>middle-income countries”,<br>differentiated service<br>delivery” |

**Figure 1.**
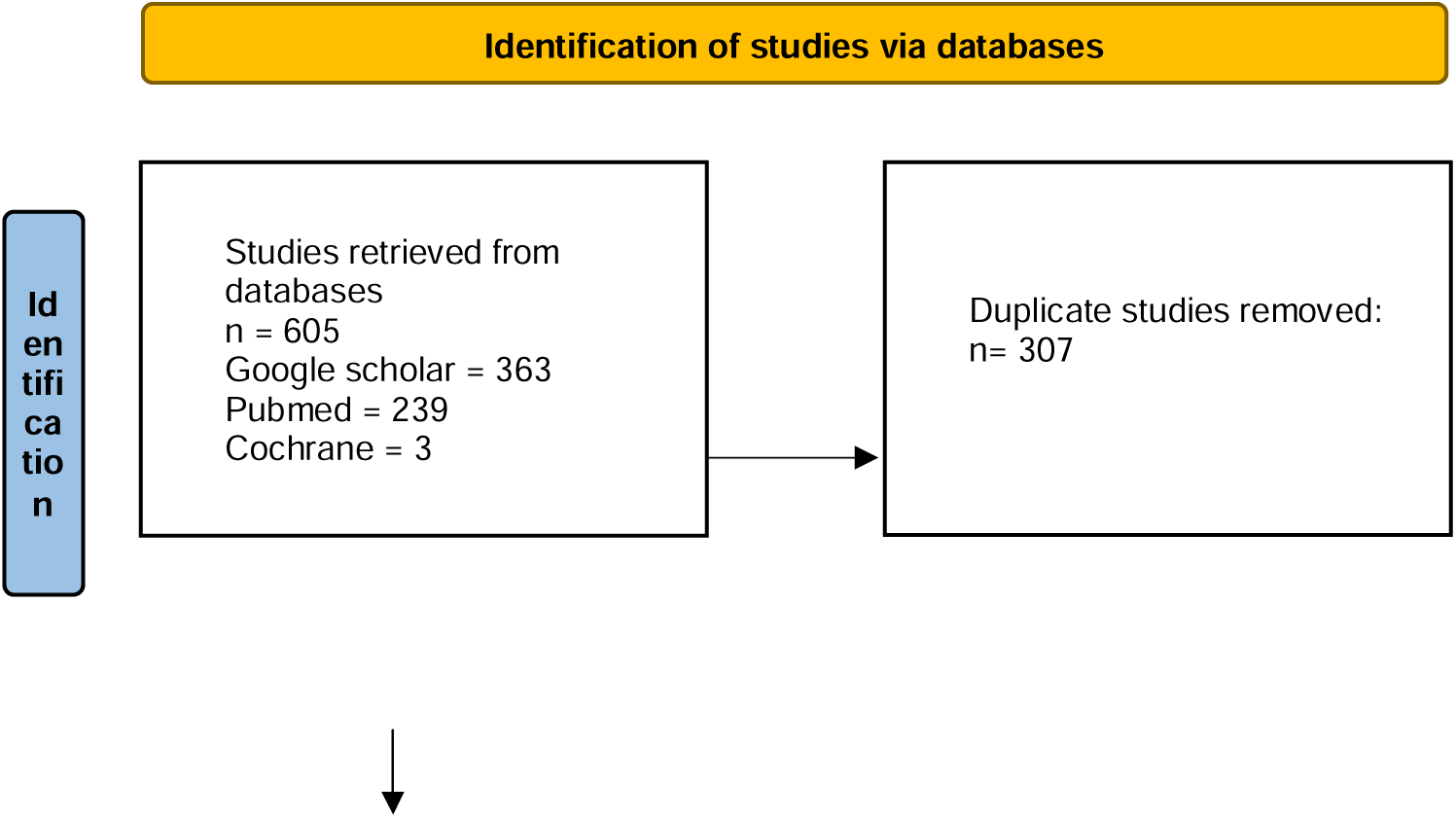
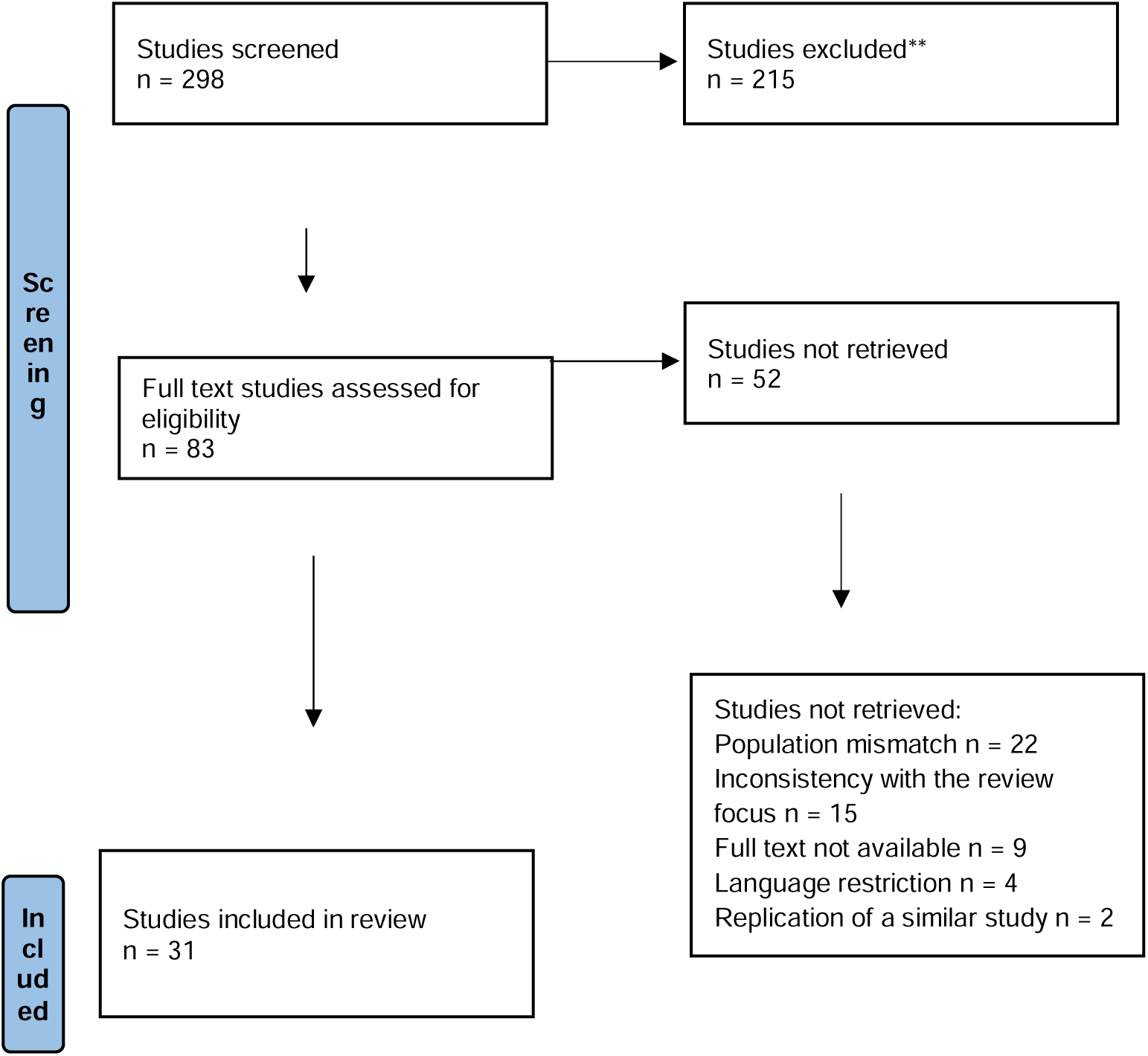
PRISMA Flow Diagram showing number of studies retrieved, screened and included in the review.

Using the PCC guide a comprehensive search was conducted online to identify relevant studies, and reports with required information based on the guide. Screening and selection criteria were clearly defined based on these PCC elements, ensuring a focus on studies that provide data on keywords such as “HIV care,” linkage to care”, “retention to care”, “case management,” “clinician partnership,” “interprofessional collaboration”, “team-based care”, “patient engagement”, “community health services” and “treatment adherence.” Data extraction involved gathering detailed information on these keywords. Synthesis of the findings was used to map the existing evidence, identify gaps in knowledge, and provide recommendations for practice and future research.

### Inclusion Criteria

1. Peer-reviewed studies and reports that provide data on collaboration between case managers and HIV clinicians, interventions aimed at improving care continuum, linkage, retention in HIV care, adherence to ART, and evaluations of collaborative models in diverse healthcare settings.
2. Studies that focus on Sub-Saharan Africa or with clear policies that reflect or (and) could aid the Sub-Saharan African countries.
3. Studies that were conducted within the last 10 years (from 2014 – 2024) and reflect current practices and outcomes.

### Exclusion Criteria

1. Studies that do not focus on HIV care, those without clear provision on interprofessional partnerships that improve care continuum.
2. Articles not available in English.
3. Studies that were conducted more than 10 years ago.

### Study Selection

The study selection was guided by the eligibility criteria that align with the aim and objectives of the scoping review. Typically, studies were selected based on their relevance to the research questions, research quality, and context-specific insights into the Nigerian healthcare setting. The selection process involved screening titles and abstracts followed by full-text review to ensure that the studies met the predefined inclusion criteria. Studies that did not meet the inclusion criteria or that did not provide clear data on the PCC elements were excluded to maintain the focus and quality of the review.

## Results

### Data Extraction Table on the Review Conducted on the Topic “Establishing effective partnerships among case managers and clinicians to improve linkage to and engagement in HIV care: A scoping review”

A total of 31 studies were used for this scoping review based on the established inclusion and exclusion criteria with a major focus on high and low resource settings. The table below shows data extraction summary for ten of the studies that are most closely related to the research question and key themes. These papers provide a geographic and contextual representation of the review and also interventions and findings that can significantly contribute to HIV care continuum.

| Author and year of the Publication | Country of Publication | Aim of the Research | Methodology and Sample Size | Result | Recommendations |
| --- | --- | --- | --- | --- | --- |
| Vorkoper et al. (2022) | United States | To analyze the use of implementation science in HIV prevention and treatment interventions for adolescents and young adults (AYA) in Sub-Saharan Africa (SSA). | A scoping review based on PRISMA-ScR guidelines, analyzing 44 studies conducted in 13 SSA countries. Data were assessed for implementation science outcomes such as acceptability, feasibility, and fidelity. | The review reported that most studies focused on prevention (61.4%) with minimal focus on care (6.8%). Common implementation outcomes were acceptability (25.8%) and feasibility (43.2%). Studies rarely used established implementation frameworks. | To enhance study designs, integrate established implementation science frameworks, and expand focus across the HIV care continuum. |
| Meya et al. (2022) | Uganda | The aim of the research was to evaluate the effect of case management on HIV treatment retention and viral suppression among key populations in Kalangala District, Uganda. | A quasi-experimental pre-post study compared 6-month retention and viral suppression before and after implementing a case management intervention involving facility and community case managers. | The study found that retention improved from 73% to 83% post-intervention, with significant gains among mobile populations like fisherfolk. Viral suppression rates remained constant (~87%). | The study recommended scaling up tailored case management to improve linkage and retention for hard-to-reach populations. |
| Besada et al. (2018) | South Africa | The research aimed to examine the roles of community cadres in enhancing retention in prevention of mother-to-child transmission (PMTCT) services across four African | The study employed a qualitative rapid appraisal through interviews and focus groups with stakeholders in Malawi, Uganda, Côte d'Ivoire, and Democratic Republic of Congo (DRC). | The study showed that community cadres improved linkage, adherence, and retention by conducting household mapping, demand creation, and follow-ups. Their roles varied | The study recommended standardization of and investment in community cadre systems to strengthen health service delivery. |
|  |  | countries. |  | significantly across countries. |  |
| Chinyandura et al. (2022) | South Africa | The aim of the research was to evaluate the effectiveness of a holistic, patient-centered case management model in improving retention in HIV care and adherence to ART among PLHIV in Johannesburg, South Africa. | The study employed a qualitative evaluation design using semi-structured interviews with key informants (retention officers and social auxiliary workers) and patients. A total of 60 patients and 17 key informants participated. Participants were recruited from eight health facilities in Johannesburg District, selected based on their performance in retaining patients in care. | The study highlighted the importance of a holistic, patient-centered approach in HIV care, addressing clinical, socioeconomic, and psychosocial needs. Supportive counseling also facilitated HIV status disclosure and reduced stigma, with referrals to social auxiliary workers for severe cases. | The study recommended the adoption of a holistic care model where health systems transition towards patient-centered care models that integrate clinical care with socioeconomic and psychological support, and ongoing training for case managers. |
| Shamu et al. (2019) | South Africa | The research aimed to evaluate the outcomes of a collaborative | The study utilized a mixed-methods approach, combining routine | The study found that linkage rates improved dramatically from 27% to | The study suggested addressing the health system inefficiencies, reduce client |
|  |  | intervention to improve linkage to HIV care in South Africa. | programmatic data with interviews of program managers, implementers, and clients. | 85% over two years, aided by client escort services, task shifting, and enhanced teamwork. | burden, and foster collaborative partnerships for sustainable outcomes |
| Comins et al. (2024) | South Africa | The aim of the research was to identify determinants of viral suppression among female sex workers (FSW) living with HIV in Durban, South Africa. | The study used baseline data analysis from the Siyaphambili study, focusing on individual, network, environmental, and structural factors impacting viral suppression. | The result showed that only 38% achieved viral suppression, with key barriers being housing instability, stigma, and substance use. Older age and hormonal contraception use were positively associated with suppression. | The authors recommended targeted interventions to younger FSW and address structural issues like housing and stigma. |
| Rachlis et al. (2016) | Kenya | The aim of the research was to explore barriers and facilitators affecting linkage and retention in care for HIV, TB, | A qualitative study was employed using in-depth interviews and focus groups with 235 participants, including patients, caregivers, | The study found that barriers included stigma, logistical challenges, lack of knowledge, and cultural beliefs, while facilitators included | The study recommended the Strengthening of patient-provider interactions and address structural and personal barriers to care through tailored interventions. |
|  |  | and hypertension under the AMPATH program in Kenya. | community leaders, and healthcare providers. | supportive patient-provider relationships and social support networks. |  |
| Keane et al. (2016) | United States | The research aimed to review interventions mitigating loss to follow-up (LTFU) at various HIV care continuum stages in sub-Saharan Africa. | The study involved a systematic review of 13 studies, assessing interventions like community support, point-of-care CD4 testing, and ART distribution models. | The result indicated that community-supported programs and point-of-care testing significantly improved linkage and retention. LTFU rates decreased in innovative intervention settings. | The article suggested comprehensive, phased approaches addressing structural and individual barriers across the care continuum. |
| Williams et al. (2021) | Eswatini | The research aimed to evaluate the impact of a community-based HIV testing and linkage program on ART initiation and retention in Eswatini. | The methodology involved case management including peer-delivered services, psychosocial support, and treatment navigation, with tailored interventions in two phases. | The research reported increased HIV testing among family members and associates. ART initiation rates improved, with 95% of identified HIV-positive individuals starting treatment. | Based on the findings, the research recommended expanding case management to high-risk contacts and integrating psychosocial support with treatment navigation was proposed to sustain outcomes |
| Masquillier et al. (2016) | South Africa | The research aimed to examine how HIV/AIDS competence within households influences the effectiveness of community-based treatment adherence support for PLHIV in South Africa. | The study utilized a combination of qualitative research methods, including participatory observations, in-depth interviews, and focus group discussions. The fieldwork was conducted in townships on the outskirts of Cape Town, South Africa | The research found that the effectiveness of community-based treatment adherence support is significantly influenced by the level of HIV/AIDS competence within the household. Households with high competence levels are more supportive and facilitate better patient outcomes. Community health workers (CHWs) play a crucial role in bridging the gap between healthcare services and patients, particularly in resource-limited settings. | The article recommended designing community-based adherence support programs that consider the household context and dynamics to address specific barriers and leverage supportive household environments. Also providing a comprehensive training for CHWs to navigate different household dynamics and reduce stigma associated with their visits. |

### Thematic Analysis of Result Findings

The following themes were prominent in the reviewed studies:

### Collaboration and Role Clarity

One of the predominant themes in improving linkage to and engagement in HIV care is the importance of collaboration and role clarity between case managers and clinicians. Studies emphasize that a lack of clear role definitions often leads to confusion and overlaps, undermining teamwork and the continuity of care (Rahman et al., 2021). Sanchez et al. (2022) reported the importance of CHWs and Case Managers being integrated within clinical teams with clear role definitions and expectations to avoid overlap and ensure cohesive teamwork. The presence of fragmentation can impede the development of integrated care plans and result in missed opportunities to address patients’ comprehensive needs (Shamu et al., 2019).

Clinicians, including physicians, nurse practitioners, and physician assistants, provide direct medical care, including diagnosis and ART management. On the other hand, case managers (often non-clinical professionals) focus on coordinating care services to ensure patients remain engaged in the HIV care continuum (Khalili & Landovitz, 2020; Besada et al., 2018). Effective partnerships hinge on defining these roles clearly while fostering mutual respect and understanding of each other’s contributions to patient care.

### Policy and Training Gaps

The lack of standardized protocols and insufficient training are recurring themes in the studies reviewed. The need for ongoing professional development to equip both clinicians and case managers with the skills necessary for effective collaboration has been highlighted by several studies (Besada et al., 2018; Wijesinghe & Alexander, 2020). Patient-provider relationships are strengthened through training programs that emphasize on interprofessional collaboration, communication, cultural competency, and support for case managers, thereby addressing gaps and enhancing care delivery (Chinyandura et al., 2022).

### Patient-Centered Approach

Collaborative partnerships have a demonstrable impact on patient outcomes, particularly in improving linkage to care, retention, and ART adherence. For instance, interventions that integrate case management with clinical care have shown significant improvements in retention rates, particularly among marginalized and mobile populations such as fisherfolk in Uganda (Meya et al., 2022). As reported by Tolley et al., (2022) and Chinyandura et al., (2022), strong, supportive relationships between case managers and patients facilitated better communication and support for ART adherence.

Holistic care models (incorporating social, psychological, and clinical support) have been effective in reducing stigma, improving disclosure rates, and enhancing ART adherence (Masquillier et al., 2016; Chinyandura et al., 2022). This further highlight the value of multidisciplinary approaches in achieving better health outcomes and reducing HIV transmission rates.

### Leveraging Community Resources and Interdisciplinary Approaches

Household context and dynamics are to be considered in designing community-based adherence support programs (Masquillier et al., 2016). The adoption of interdisciplinary models is critical to addressing the multifaceted needs of PLHIV. Zablotska and O’Connor, (2017) encouraged fostering collaboration between case managers, clinicians, and community organizations to provide comprehensive care through joint training sessions and shared goals. Studies by Meya et al. (2022) and Williams et al. (2021) have highlighted the success of team-based care in Sub-Saharan Africa, where case managers work alongside clinicians and CHWs to improve ART adherence and retention rates.

### Addressing Barriers

Numerous structural and systemic barriers inhibit effective collaboration between case managers and clinicians. High caseloads, inadequate staffing, and insufficient training are pervasive issues that limit the capacity of both groups to engage fully in collaborative care (Rahman et al., 2021; Fuller et al., 2024). Differences in remuneration and status between case managers and clinicians also create tensions that can undermine teamwork (Shamu et al., 2019).

Stigma, social welfare and discrimination remain significant challenges in many healthcare settings, particularly in low- and middle-income countries. These factors discourage PLHIV from seeking care and reduce engagement in treatment programs (Rachlis et al., 2016; Comins et al., 2024). Okeke et al., (2014) suggested adopting decentralization and task-shifting strategies in resource-poor settings to improve access and retention in care. It is optimal that efforts are increased to provide resources, foster cultural competency, ensure equitable treatment across care teams and disseminate accurate and actionable healthcare information through trusted sources. Addressing non-clinical barriers such as housing and transportation and leveraging high levels of trust in HIV clinicians to educate patients about healthcare policies, are shown to improve engagement in care (Kaperak et al., 2020).

## DISCUSSION

This study explored and identified effective strategies for establishing effective partnerships between case managers and clinicians in enhancing the HIV care continuum. These partnerships, which integrate medical and non-medical support services, address multifaceted challenges faced by PLHIV especially KP, including stigma, socioeconomic barriers, and psychological distress (Keane et al., 2016; Vorkoper et al., 2023). Well-coordinated, interdisciplinary approaches have consistently been shown to lead to improved retention, ART adherence, and overall health outcomes (Chinyandura et al., 2022; Brennan-Ing et al., 2016).

However, systemic barriers persist, such as resource limitations, medical hierarchy, unclear role definitions, and heavy workloads. Resource-constrained settings, in particular, report high caseloads and inadequate staffing, which limit the ability of care teams to provide tailored interventions (Rahman et al., 2021; Fuller et al., 2024; Sanchez et al., 2022). These barriers are similar to those identified by Wijesinghe and Alexander, (2020) and Sousa et al., (2024), who reported that stigma, lack of resources, inadequate training of case managers, discrepancies in patient records and insufficient clinical training for community case managers were identified as key challenges in the United States and Mozambique respectively. Addressing these challenges requires significant investment in healthcare infrastructure, staffing, and training.

Community engagement has emerged as a critical factor in bridging gaps between healthcare providers and patients, particularly in rural and underserved areas (Masquillier et al., 2016; Williams et al., 2021). However, effective strategies vary across different geographic and socioeconomic settings as was reported by Meya et al. (2022) and Chinyandura et al., (2022), in resource-poor or socioeconomically disadvantaged settings, strategies such as decentralization of HIV care services, task-shifting and addressing barriers such as poverty and unemployment have proven effective in improving retention in care while in highly resourced settings, strength-based case management and peer navigation are more commonly implemented and have shown significant positive outcomes. Equally, studies such as Alderwick et al. (2021) challenge the assumption that inter-organizational collaboration inherently leads to better outcomes, emphasizing the importance of contextualized strategies that have shown inherent success rate. These submissions agree with those made by Agaba et al., (2018) who reported that different settings, including geographic and socio-economic contexts, significantly affect the effectiveness of these partnerships. Rural areas may face challenges related to accessibility and resource availability, whereas urban settings might deal with higher patient volumes and more complex social determinants of health.

Patient-centered care models address both clinical and socioeconomic needs. Chinyandura et al. (2022) advocate for such models, which have shown to improve ART adherence and reduce stigma. However, Comins et al., 2024 argues that structural inequities, including housing instability and stigma, still remain significant barriers and integrating patient-centered approaches with structural interventions is essential for sustainable outcomes. Similarly, psychological support, individualized care, tailored to patients’ preferences and needs, correlates with higher CD4 counts and lower viral loads (Sarfo et al., 2017; Eshiet & Njoku, 2023; Sousa et al., 2024). Okeke et al. (2014) however, caution that such interventions must account for resource constraints, as one-size-fits-all models may not be sustainable.

There is ongoing debate about the relative effectiveness of micro-level (individual-focused) and macro-level (system-focused) strategies. Keane et al. (2016) argue for broader structural interventions, such as point-of-care testing and community-supported ART distribution models, while Brennan-Ing et al. (2016) emphasize the indispensability of comprehensive case management that encompasses psychosocial, medical, and logistical support. This suggest that integrating both approaches may yield optimal results.

Fostering trust and mutual respect between case managers and clinicians is fundamental to building effective partnerships. Training programs should emphasize teamwork, communication, and the importance of interprofessional collaboration (Besada et al., 2018; Shamu et al., 2019). Flexible, community-based case management models, as proposed by Williams et al. (2021), have demonstrated high ART initiation and retention rates, further supporting the need for adaptable frameworks. Together, these perspectives reinforce the interconnectedness of social, structural, and clinical factors in effective HIV care.

## CONCLUSION

All the studies reviewed emphasized the importance of effective partnerships between case managers and HIV clinicians which are vital for improving linkage to and retention in HIV care (Rahman et al., 2021). The integration of various models, such as strength-based case management, peer navigation, and holistic, patient-centered approaches, has shown significant benefits in enhancing patient engagement and health outcomes (Sanchez et al., 2022; Chinyandura et al., 2022; Brennan-Ing et al., 2016). Additionally, focusing on the dynamic nature of patient engagement and the importance of active self-management is crucial (Agaba et al., 2018; Keene et al., 2023).

## RECOMMENDATION

Based on the findings of this study, it is essential that hospitals or clinics employ a patient-centered approach, adopting holistic care models that integrate clinical, socioeconomic, and psychological support tailored to address the comprehensive needs of PLHIV. There is need for integration and role clarity and ensuring consistent and clear communication between all team members and the provision of comprehensive training and support to enhance the skills and confidence of these HIV clinicians and case managers. Implementing strategies to overcome barriers such as stigma, housing instability, and lack of access to technology is also found to be crucial in improving linkage and retention in HIV care. Develop policies to reduce stigma, improve access to technology, and provide financial support for housing and transportation. It is also important to leverage community resources by collaborating with community organizations and advocating for policies to improve healthcare access and reduce costs. Finally, it is paramount to regularly evaluate the effectiveness of implemented interventions and make necessary adaptations based on patient feedback and emerging best practices.

## Data Availability

All data produced in the present work are contained in the manuscript

## REFERENCES

Agaba, P.A., Meloni, S.T., Agbaji, O.O., et al. (2018) ‘Retention in differentiated care: multiple measures analysis for a decentralized HIV care and treatment program in North Central Nigeria’, Journal of AIDS & Clinical Research, 9(2). DOI: 10.4172/2155-6113.1000756.

Alderwick, H., Hutchings, A., Briggs, A. and Mays, N. (2021) ‘The impacts of collaboration between local health care and non-health care organizations and factors shaping how they work: a systematic review of reviews’, BMC Public Health, 21, pp.1–16.

Besada, D., Goga, A., Daviaud, E., et al. (2018) ‘Roles played by community cadres to support retention in PMTCT Option B+ in four African countries: a qualitative rapid appraisal’, BMJ Open, 8(3), p.e020754. DOI: 10.1136/bmjopen-2017-020754.

Brennan-Ing, M., Seidel, L., Ansell, P., et al. (2016) ‘The impact of comprehensive case management on HIV client outcomes’, PLoS One, 11(2), p.e0148865. DOI: 10.1371/journal.pone.0148865.

Chinyandura, C., Ngoma, M., Mucheto, P., et al. (2022) ‘Supporting retention in HIV care through a holistic, patient-centred approach: a qualitative evaluation’, BMC Psychology, 10(1), p.17. DOI: 10.1186/s40359-022-00722-x.

Comins, C.A., Baral, S., Mcingana, M., et al. (2024) ‘ART coverage and viral suppression among female sex workers living with HIV in eThekwini, South Africa: Baseline findings from the Siyaphambili study’, PLOS Global Public Health, 4(5), p.e0002783. DOI: 10.1371/journal.pgph.0002783.

Eshiet, U.I. and Njoku, C.R. (2023) ‘Individualized patient care and clinical outcomes in HIV/AIDS management’, American Journal of Pharmacotherapy and Pharmaceutical Sciences, 2.

Fuller, S.M., Arnold, E.A., Xavier, J., et al. (2024) ‘Integrating community health workers into HIV care clinics: a qualitative study with health system leaders and clinicians in the Southern United States’, BMC Health Services Research, 24(1), pp.1–13.

Kaperak, C., Marks, J., Smith, A., et al. (2020) ‘A CrosslJSectional Study on the Affordable Care Act from the Perspective of People Living with HIV: The Interplay between Knowledge, Stigma, Trust, and Attitudes’, AIDS Research and Treatment, 2020(1), p.6081721. DOI: 10.1155/2020/6081721.

Keane, J., Pharr, J.R., Buttner, M.P. and Ezeanolue, E.E. (2017) ‘Interventions to reduce loss to follow-up during all stages of the HIV care continuum in sub-Saharan Africa: a systematic review’, AIDS and Behavior, 21(6), pp.1745–1754. DOI: 10.1007/s10461-016-1532-y.

Keene, C.M., Carter, C., Okello, R., et al. (2023) ‘Conceptualising engagement with HIV care for people on treatment: the Indicators of HIV Care and AntiRetroviral Engagement (InCARE) Framework’, BMC Health Services Research, 23(1), p.435. DOI: 10.1186/s12913-023-09433-4.

Khalili, J. and Landovitz, R.J. (2020) ‘HIV preexposure prophylaxis—the role of primary care clinicians in ending the HIV epidemic’, JAMA Internal Medicine, 180(1), pp.126–130. DOI: 10.1001/jamainternmed.2019.5456.

Lambdin, B.H., Cheng, B., Peter, T., et al. (2015) ‘Implementing implementation science: an approach for HIV prevention, care and treatment programs’, Current HIV Research, 13(3), pp.244–246. DOI: 10.2174/1570162X13666150504163201.

Masquillier, C., Wouters, E., Mortelmans, D., et al. (2016) ‘HIV/AIDS competent households: interaction between a health-enabling environment and community-based treatment adherence support for people living with HIV/AIDS in South Africa’, PLoS One, 11(3), p.e0151379. DOI: 10.1371/journal.pone.0151379.

Meya, D.B., Kiragga, A.N., Nalintya, E., et al. (2022) ‘Impact of an intensive facility-community case management intervention on 6-month HIV outcomes among select key and priority populations in Uganda’, AIDS Research and Therapy, 19(1), p.62. DOI: 10.1186/s12981-022-00486-9.

Mwai, G.W., Mburu, G., Torpey, K., et al. (2013) ‘Role and outcomes of community health workers in HIV care in sublJSaharan Africa: a systematic review’, African Journal of Reproductive and Gynaecological Endoscopy, 16(1). DOI: 10.7448/IAS.16.1.18586.

Okeke, N.L., Ostermann, J. and Thielman, N.M. (2014) ‘Enhancing linkage and retention in HIV care: a review of interventions for highly resourced and resource-poor settings’, Current HIV/AIDS Reports, 11, pp.376–392. DOI: 10.1007/s11904-014-0233-9.

Øgård-Repål, A., Berg, R.C. and Fossum, M. (2023) ‘Peer support for people living with HIV: A scoping review’, Health Promotion Practice, 24(1), pp.172–190. DOI: 10.1177/15248399221119712.

Peters, M.D.J., Godfrey, C.M., McInerney, H., et al. (2015) ‘Guidance for conducting systematic scoping reviews’, International Journal of Evidence-Based Healthcare, 13(3), pp.141–146.

Rachlis, B., Naanyu, V., Wachira, J., et al. (2016) ‘Identifying common barriers and facilitators to linkage and retention in chronic disease care in western Kenya’, BMC Public Health, 16, pp.1–15. DOI: 10.1186/s12889-016-3787-0.

Rahman, R., Pinto, R.M. and Troost, J.P. (2021) ‘Examining Interprofessional Collaboration across case managers, peer educators, and counselors in New York City’, Social Work in Public Health, 36(4), pp.448–459. DOI: 10.1080/19371918.2021.1905131.

Sanchez, A.L., Pinto, R.M., Rahman, R., et al. (2022) ‘Stakeholder perspectives on MAPS’, JAIDS Journal of Acquired Immune Deficiency Syndromes, 90(S1), pp.S190–S196. DOI: 10.1097/QAI.0000000000002979.

Santos, W.M.D., Secoli, S.R. and Püschel, V.A.D.A. (2018) ‘The Joanna Briggs Institute approach for systematic reviews’, Revista Latino-Americana de Enfermagem, 26:3074.

Sarfo, B., Antwi, K., Boafo, J., et al. (2017) ‘HIV case management support service is associated with improved CD4 counts of patients receiving Care at the Antiretroviral Clinic of Pantang hospital, Ghana’, AIDS Research and Treatment, 2017(1), p.4697473. DOI: 10.1155/2017/4697473.

Shamu, S., Slabbert, J., Guloba, G., et al. (2019) ‘Linkage to care of HIV positive clients in a community based HIV counselling and testing programme: a success story of non-governmental organisations in a South African district’, PLoS One, 14(1), p.e0210826. DOI: 10.1371/journal.pone.0210826.

Sousa, B., Mate, K.S., Gimbel, S., et al. (2024) ‘Adopting Data to Care to Identify and Address Gaps in Services for Children and Adolescents Living With HIV in Mozambique’, Global Health: Science and Practice, 12(2). DOI: 10.9745/GHSP-D-23-00130.

Suleiman, I.A. and Momo, A. (2016) ‘Adherence to antiretroviral therapy and its determinants among persons living with HIV/AIDS in Bayelsa state, Nigeria’, Pharmacy Practice, 14(1). DOI: 10.18549/PharmPract.2016.01.631.

Tolley, E.E., Johnston, C.C., Milkovich, K., et al. (2022) ‘The role of case management in HIV treatment adherence: HPTN 078’, AIDS and Behavior, 26(9), pp.3119–3130. DOI: 10.1007/s10461-022-03644-2.

Vorkoper, S., Tahlil, K.M., Sam-Agudu, N.A., et al. (2023) ‘Implementation science for the prevention and treatment of HIV among adolescents and young adults in sub-Saharan Africa: a scoping review’, AIDS and Behavior, 27(Suppl 1), pp.7–23. DOI: 10.1007/s10461-022-03780-1.

Walker, D., Moucheraud, C., Butler, D., et al. (2023) ‘Experiences with telemedicine for HIV care in two federally qualified health centers in Los Angeles: a qualitative study’, BMC Health Services Research, 23(1), p.156. DOI: 10.1186/s12913-023-09107-1.

WHO (World Health Organization) (2024) HIV https://www.who.int/data/gho/data/themes/hivaids#:~:text=Global%20situation%20and%20trends%3A,at%20the%20end%20of%202022, accessed 17 June 2024.

Wijesinghe, S. and Alexander, J.L. (2020) ‘Management and treatment of HIV: are primary care clinicians prepared for their new role?’, BMC Family Practice, 21, pp.1–11. DOI: 10.1186/s12875-020-01198-7.

Williams, D., MacKellar, D., Dlamini, M., et al. (2021) ‘HIV testing and ART initiation among partners, family members, and high-risk associates of index clients participating in the CommLink linkage case management program, Eswatini, 2016–2018’, PLoS One, 16(12), p.e0261605. DOI: 10.1371/journal.pone.0261605.

Zablotska, I.B. and O’Connor, C.C. (2017) ‘Preexposure prophylaxis of HIV infection: the role of clinical practices in ending the HIV epidemic’, Current HIV/AIDS Reports, 14, pp.201–210. DOI: 10.1007/s11904-017-0367-7.

